# Looking under the lamp-post: quantifying the performance of contact tracing in the United States during the SARS-CoV-2 pandemic

**DOI:** 10.1101/2023.03.27.23287812

**Authors:** Henry Bayly, Madison Stoddard, Debra Van Egeren, Eleanor J Murray, Julia Raifman, Arijit Chakravarty, Laura F White

**Affiliations:** Department of Biostatistics, Boston University School of Public Health, Boston, MA, USA; Fractal Therapeutics, Cambridge, MA, USA; Stanford University, Palo Alto, CA, USA; Department of Epidemiology, Boston University School of Public Health, Boston, MA, USA; Department of Health Law, Policy and Management, Boston University School of Public Health, Boston, MA, USA

## Abstract

Contact tracing forms a crucial part of the public-health toolbox in mitigating and understanding emergent pathogens and nascent disease outbreaks. Contact tracing in the United States was conducted during the pre-Omicron phase of the ongoing COVID-19 pandemic. This tracing relied on voluntary reporting and responses, often using rapid antigen tests (with a high false negative rate) due to lack of accessibility to PCR tests. These limitations, combined with SARS-CoV-2’s propensity for asymptomatic transmission, raise the question “how reliable was contact tracing for COVID-19 in the United States”? We answered this question using a Markov model to examine the efficiency with which transmission could be detected based on the design and response rates of contact tracing studies in the United States. Our results suggest that contact tracing protocols in the U.S. are unlikely to have identified more than 1.65% (95% uncertainty interval: 1.62%-1.68%) of transmission events with PCR testing and 0.88% (95% uncertainty interval 0.86%-0.89%) with rapid antigen testing. When considering an optimal scenario, based on compliance rates in East Asia with PCR testing, this increases to 62.7% (95% uncertainty interval: 62.6%-62.8%). These findings highlight the limitations in interpretability for studies of SARS-CoV-2 disease spread based on U.S. contact tracing and underscore the vulnerability of the population to future disease outbreaks, for SARS-CoV-2 and other pathogens.

## Introduction

The management and control of infectious disease has been a signal modern achievement. Advances in epidemiological techniques pioneered during the 19^th^ century established public health as a discipline. Overlapping with, but distinct from the medical establishment and the biopharmaceutical industry, modern public health organizations have sought to control disease using nonpharmaceutical interventions (NPIs).

Contact tracing is a cornerstone of the public-health response, particularly with emergent pathogens and nascent disease outbreaks [1]. Effective contact tracing facilitates estimates of epidemiological parameters describing disease spread. In the current COVID-19 pandemic, rigorous early studies relying on contact tracing revealed key epidemiological features of SARS-CoV-2 such as asymptomatic transmission [2,3], superspreading [4], and aerosol transmission [5–9]. This provided a basis for projecting the future course of the outbreak and designing a public health response.

Effective contact tracing is also critical for limiting onward spread through the deployment of test-and-trace and isolation protocols. Many Asia-Pacific countries effectively limited SARS-CoV-2 community spread for the first two years of the pandemic, relying on contact tracing with isolation of contacts, including strict testing and isolation efforts at their borders. For example, South Korea used methods such as tracking credit card transactions and using closed circuit televisions to link contacts together [10]. In China, specifically Hubei, suspected contacts were placed under monitored house arrest throughout their quarantine period [11]. This strategy permitted high levels of withing country contact and mobility while keeping case counts low [12–18].

In the U.S., contact tracing was primarily performed in the pre-Omicron era, and largely abandoned in early 2022 [19]. It has been widely recognized that contact tracing in the U.S. has not slowed disease transmission [20]. Part of the challenge has been that the process varied from state to state and relied on individual initiative and access to testing [19]. This meant that an individual typically must be symptomatic, voluntarily seek testing, and have their positive result reported to initiate contact tracing [21]. Public health officials initiated an investigation by asking the index case to identify their contacts, who in turn would be interviewed. The exposed contacts were monitored for symptoms and could choose to test for SARS-CoV-2 five days after exposure. If positive, the contact (now a secondary case) would be asked to name their contacts.

The process was largely voluntary, allowing for selection bias and many missed transmission chains. There was often no system for identifying close contacts whom the index case did not know personally. Many published papers noted that many named contacts were not successfully traced [22–24] and not all symptomatic contacts were willing to undergo testing [25]. A systematic surveillance-based cross-sectional study in the U.S. showed that 2 out of 3 index cases of COVID-19 were either not reached by tracers or declined to share contacts. Only 70% of named contacts agreed to be interviewed, and only 50% of those contacts were monitored, leading to an average of less than one contact per index case being monitored [26]. Additionally, the CDC-recommended 15 minutes of contact within six feet over a 24-hour period was somewhat arbitrary and never updated, even as evidence emerged indicating that COVID-19 could be transmitted through brief interactions.

The implications of these limitations in contact tracing are significant. The relatively high reproductive number for SARS-CoV-2 [27] would suggest that many transmission chains generated from a single index case went undetected. Additionally, asymptomatic transmission and superspreading behavior would also impact the efficacy of contact tracing for infection control and the generalizability of inferences made about transmission dynamics [28,29].

In keeping with this voluntary and symptom-gated approach to contact tracing, there are many examples of minimally observed onward transmission in settings where transmission would be expected. This includes studies involving children with strong implications for policies related to schools. The results of studies investigating children and COVID-19 transmission have documented limited forward transmission, but this is often in context of significant mitigation strategies being in place or incomplete contact tracing [30–32]. During the initial omicron surge, when contact tracing was limited, schools struggled to remain open, reported high absenteeism rates, and in some cases, relied on the national guard to teach courses and due to incomplete contact tracing it was unclear what role children in schools played in transmission [33,34]. In one another case, two COVID-19 positive hairdressers in Missouri saw 139 clients over a ten-day period, with no reported onward transmission [24]. Notably, of the exposed clients, only ~75% (n=104) responded to contact tracers’ requests for interviews, and only ~50% (n=67) agreed to be tested. Biases in willingness to respond to interviews or agree to testing may have concealed many onward transmission events.

Another example, demonstrated the challenges in identifying both primary and secondary infections, was the Sturgis motorcycle rally in August 2020. Following this 10-day event in Meade County, South Dakota (attended by approximately 460,000 persons [35] without [36–38] any mask-wearing requirements or other mitigating policies [39]), there was a wave of COVID-19 cases in Meade County and South Dakota. The counties outside of South Dakota that contributed the highest inflows of rally attendees experienced a 6.4-12.5% increase in COVID-19 cases relative to counties without inflows [40]. Despite evidence of population-level changes in COVID-19 case counts following the rally, the CDC and Minnesota Department of Health were able to identify only 21 person-to-person transmission events [41]. Out of the 86 positive cases, only 41 reported being in close contact (defined as being within 6 feet of another person for ≥15 minutes) with other people, and they reported an average of 2.5 close contacts. Both statistics are implausible for a 10-day motorcycle rally featuring indoor dining and concerts [42–44]. The CDC’s report does not specify how many of the 102 secondary contacts were tested, consistent with other U.S. contact-tracing studies [45,46].

Examples such as these, coupled with the unique features of SARS-CoV-2 contact tracing in the U.S. during the early part of the pandemic raise the question “what was the efficiency of contact tracing, as it was implemented in the U.S.”? We answered this question using Markov Chain modeling to synthesize data from multiple sources of information on testing and contact tracing completeness to estimate the efficiency with which onward transmission could be detected. Our model-based approach sought to quantify two metrics of performance for contact tracing: 1) the percentage of all transmission pairs identified in a disease cluster/outbreak, and 2) the percentage of onward transmission events identified from a known index case. These metrics correspond broadly to the two contact-tracing scenarios described above, the Sturgis motorcycle rally study (seeking all transmission pairs) and the Missouri hairdressers (seeking onward transmission from a known index case). Our results suggest that, as may be expected, contact tracing protocols in the United States are unlikely to have identified more than a vanishingly small fraction of transmission events. We contrast this with a similar model run that incorporates data from Asian countries with more comprehensive contact tracing protocols in place, which yields a larger fraction of identified transmission events.

## Methods

### Model

We created a Markov Model to represent the sequence of events in contact tracing depicted in Figure 1. The model captures the steps in contact tracing beginning with identifying a primary infectious individual through testing and then engaging that person in contact tracing by accurately identifying their contacts. The final steps of the model details engaging the infected contact and completing testing. We focus on estimating the probability of identifying infected contacts only. We parameterize the model steps through literature review. We capture uncertainty in the transition parameters by sampling over a literature-informed uncertainty range of the parameters 10,000 times. When no data was available on a parameter value, we sweep over the range of [0,1].

**Figure 1.**
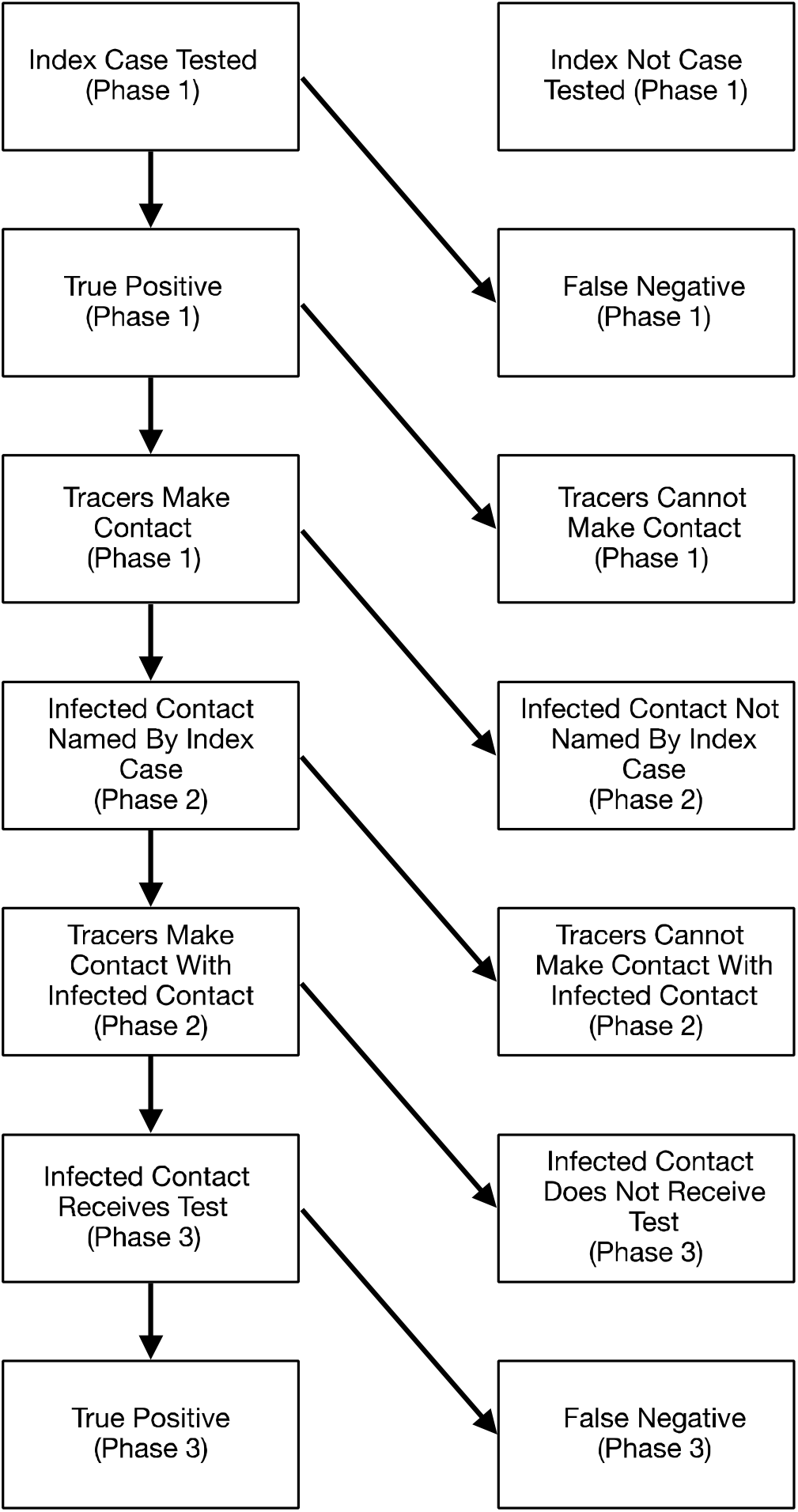
Schematic representation of steps required to correctly identify secondary cases of infected individuals. Each step is a binary variable and represents a point in the process where failure can occur. Steps 1-3 coincide with Phase 1 (identifying positive index cases). Steps 4 and 5 coincide with phase 2 (identifying contacts of positive cases). Finally, steps 6 and 7 coincide with Phase 3 (identifying positive cases among contacts).

### Literature search and model parameterization

We reviewed the literature to produce estimates for each of the parameters of our model (Figure 1). We used the search term “(covid 19 or covid-19 or covid19) AND (case investigation or contact tracing) AND (united states or US)” on PubMed to inform parameters that define the likelihood of individuals naming their close contacts and of those contacts responding to a contact tracing encounter and being tested. We collected data from contact tracing investigations that reported the proportion of positive cases that named contacts, the proportion of named contacts reached, the proportion of contacts that cooperated with tracers, and/or the proportion of contacts tested.

We also parameterized our model to represent a setting where stringent contact tracing was implemented. For this, we derived estimates from Taiwan and South Korea, which had rigorous contact tracing protocols during the initial phase of the pandemic using the search terms ‘(covid 19 or covid-19 or covid19) AND (case investigation or contact tracing) AND (taiwan)’ and ‘(covid 19 or covid-19 or covid19) AND (case investigation or contact tracing) AND (korea or south korea)’. We run the model separately for RAT and PCR tests.

The model can be divided into two distinct categories of parameters that contribute to the overall effectiveness of contact tracing: 1) efficiency of testing and 2) efficiency of contact tracing. Efficiency of testing includes the proportion of symptomatic people receiving testing, test sensitivity (RAT or PCR) [47,48], and the proportion of contacts tested. Efficiency of contact tracing aggregates the probabilities that tracers contact a positive index case, a positive index case names contacts, and named contacts are traced. Both aggregate parameters take values between [0,1]. We plot all possible combinations of these parameters on a heatmap and identify the estimates obtained for the U.S.-based and ideal scenarios.

### Code Availability

The Markov Model was implemented in Python, and code for running the simulations and plotting the results are available in a Jupyter notebook on Github (https://github.com/Henry-Bayly/ContactTracingMarkovModel).

## Results

### Literature search

Our literature search of U.S. studies yielded 1,355 papers. Of these, the first 350 were reviewed to represent a random sample of the total papers found, as our goal was to conduct a representative and not comprehensive literature review. We excluded 325 papers that did not contain data relevant to the parameters required for our model, leaving twenty-five papers with information on contact tracing parameters. When multiple papers had values for the same parameter, for instance the probability of a case naming contacts, we assumed that the true value was uniformly distributed in the range of the reported values across the papers. All parameter values are shown in Table 1; sources of the parameters are shown in Tables S1-S6. We were unable to identify precise parameter estimates for the probability of a symptomatic person receiving testing and assume a uniform distribution over [0,1].

**Table 1.**
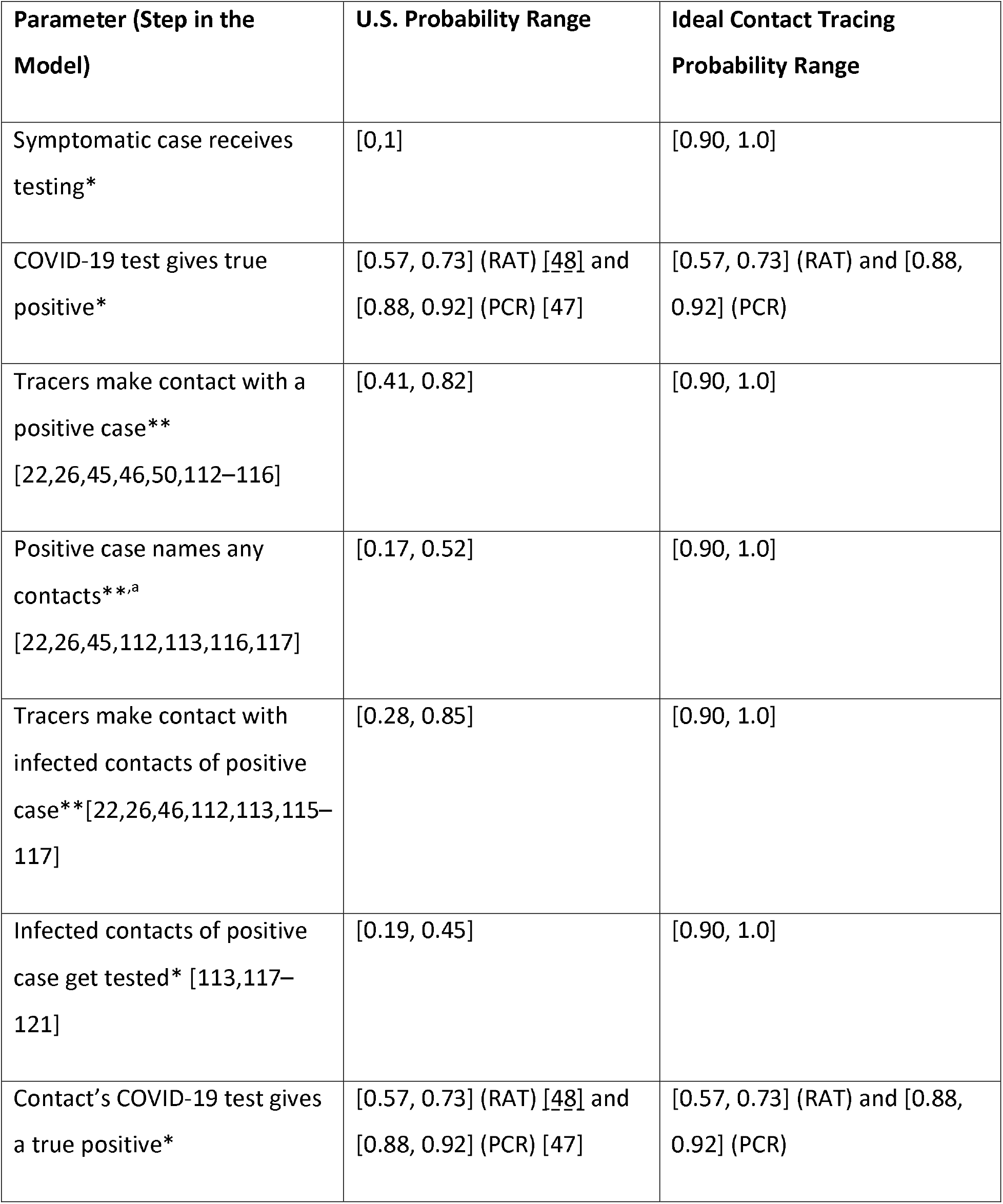
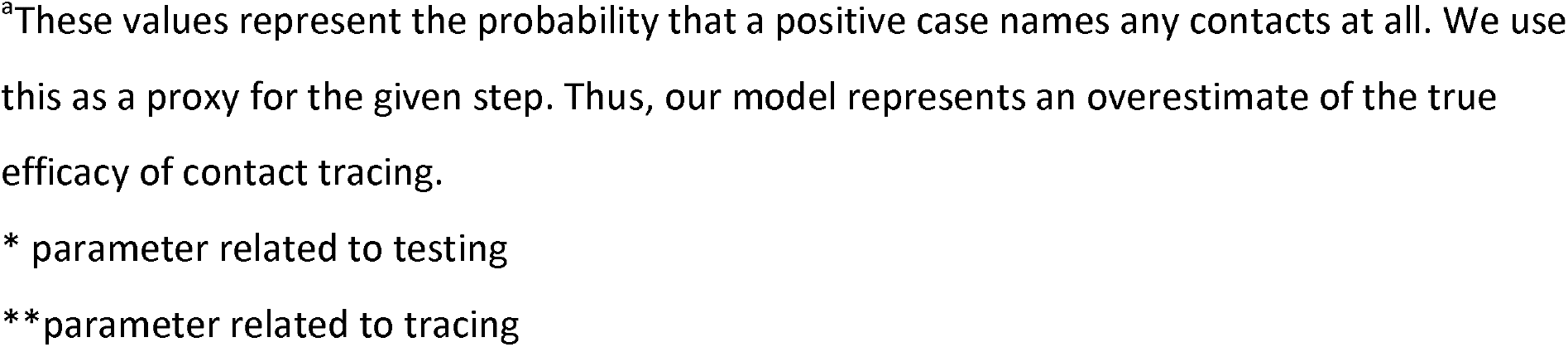
Table of parameters derived from literature that were used in the model.

| Parameter (Step in the Model) | U.S. Probability Range | Ideal Contact Tracing Probability Range |
| --- | --- | --- |
| Symptomatic case receives testing* | [0,1] | [0.90, 1.0] |
| COVID-19 test gives true positive* | [0.57, 0.73] (RAT) [48] and [0.88, 0.92] (PCR) [47] | [0.57, 0.73] (RAT) and [0.88, 0.92] (PCR) |
| Tracers make contact with a positive case**<br>[22,26,45,46,50,112–116] | [0.41, 0.82] | [0.90, 1.0] |
| Positive case names any contacts**<sup>a</sup><br>[22,26,45,112,113,116,117] | [0.17, 0.52] | [0.90, 1.0] |
| Tracers make contact with infected contacts of positive case**[22,26,46,112,113,115–117] | [0.28, 0.85] | [0.90, 1.0] |
| Infected contacts of positive case get tested* [113,117–121] | [0.19, 0.45] | [0.90, 1.0] |
| Contact's COVID-19 test gives a true positive* | [0.57, 0.73] (RAT) [48] and [0.88, 0.92] (PCR) [47] | [0.57, 0.73] (RAT) and [0.88, 0.92] (PCR) |

<sup>a</sup>These values represent the probability that a positive case names any contacts at all. We use this as a proxy for the given step. Thus, our model represents an overestimate of the true efficacy of contact tracing.
\* parameter related to testing
\*\*parameter related to tracing

We reviewed 225 papers from South Korea and Taiwan. However, we were unable to find studies quantifying contact tracing parameters related to the completeness of tracing. This appeared to be due to a much more comprehensive approach to contact tracing in these countries, negating the need to report on these parameters since it was assumed that reporting was nearly complete. For example, South Korea used traditional shoe-leather epidemiology along with large databases (global positioning system, credit card transactions, and closed-circuit television) [10] and in one study of 5,706 index cases an average of 9.9 contacts per index case were reported [10]. Taiwan similarly reported an average of 27.61 contacts per index case [49]. This contrasts with the US where the average number of non-household contacts reported in a large study was one for every three index cases [50]. Therefore, we hypothesized a realistic range of [0.9, 1.0] for all parameters not associated with testing sensitivity in our ideal model setting.

### Estimates of contact tracing efficacy

We examined the efficiency of contact tracing along the two dimensions described previously: 1) the percentage of all transmission pairs identified in a disease cluster/outbreak, and 2) the percentage of onward transmission events identified from a known index case.

We estimate a 0.88% chance (95% uncertainty range: 0.86%-0.89%) of identifying any transmission pair in the U.S. when RAT are the primary testing modality. More specifically, we estimate a 20.2% chance of identifying a positive index case, a 20.0% chance of identifying contacts given that the index case has been identified, and a 21.2% chance of identifying positive secondary cases given the index cases and contacts were identified. Using more sensitive, but less available, PCR tests, we estimate a 1.65% (95% uncertainty range: 1.62%-1.68%) chance of identifying a transmission pair (Figure 2, Table 2). Our model estimates a 27.7% chance of identifying a positive index case, a 19.8% chance of identifying contacts given that the index case has been identified, and a 29.2% chance of identifying positive cases given that the index cases and its contacts were correctly identified. By contrast, when we use an idealized scenario, based on data from East Asia, we estimate a 62.7% (95% uncertainty range: 62.6%-62.8%) chance of identifying a given transmission pair when using PCR testing and 33.5% (95% uncertainty range: 33.4%-33.6%) when using RAT (Figure 2, Table 2).

**Table 2.** Probabilities of correctly identifying positive contacts of a given index case stratified by contact tracing style and by use of RAT/PCR tests. Values in parenthesis represent the 95% uncertainty intervals.

|  | United States |  | Ideal Contact Tracing |  |
| --- | --- | --- | --- | --- |
|  | RAT | PCR Tests | RAT | PCR Tests |
| Percent of all transmission pairs in an outbreak | 0.88% (0.86%-0.89%) | 1.65% (1.62%-1.68%) | 33.5% (33.4%-33.6%) | 62.7% (62.6%-62.8%) |
| Percent of onward transmission from a known index case | 2.62% (2.59%-2.65%) | 3.62% (3.58%-3.66%) | 53.3% (53.2%-53.4%) | 73.3% (73.2%-73.4%) |

**Figure 2.**
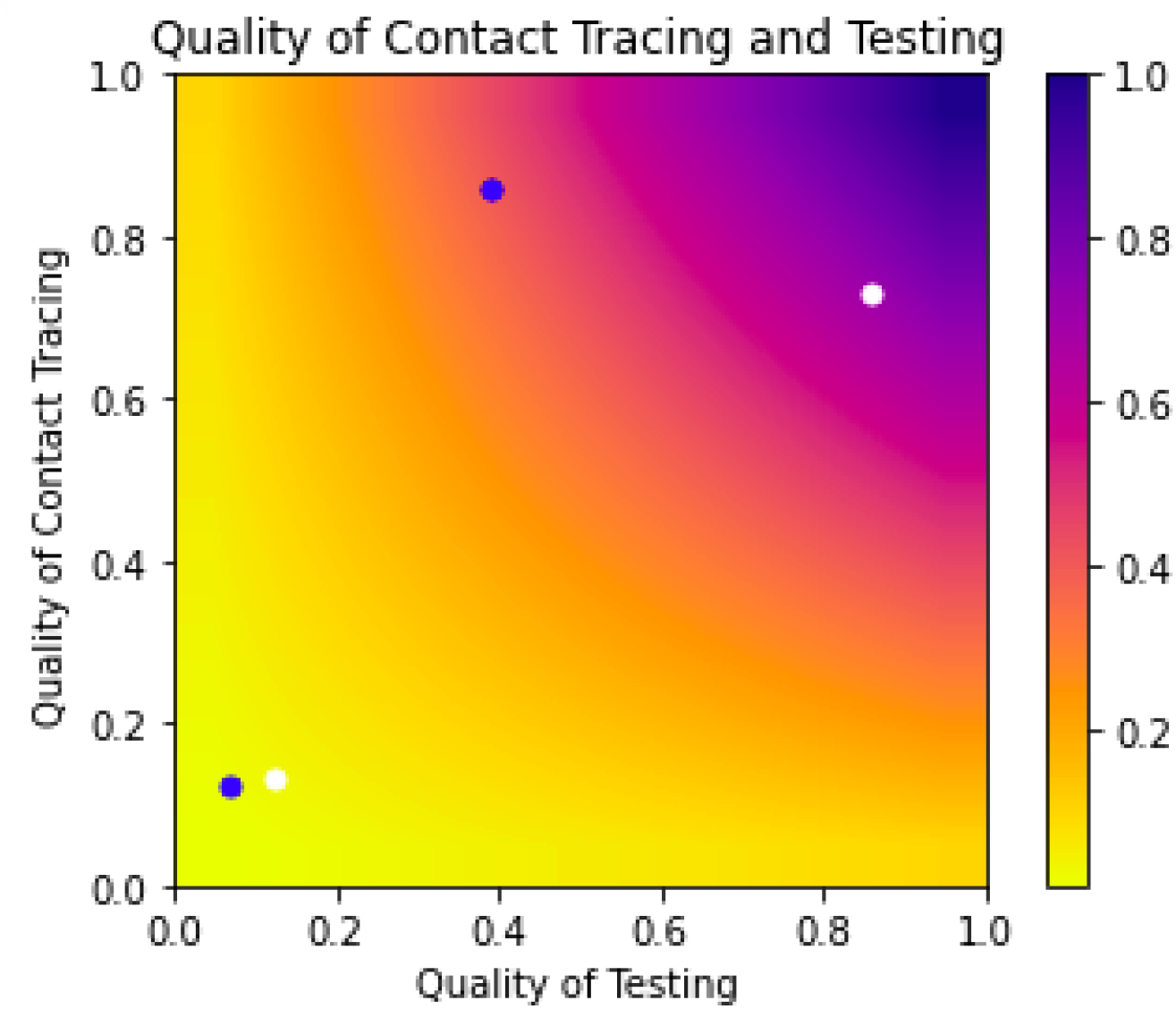
Impact of contact tracing and testing on the probability of identifying a positive contact of an infected individual with COVID-19. Quality of testing refers to parts of the process relating to testing and aggregates the following probabilities: symptomatic people receiving testing, symptomatic index cases receive true positive test result (i.e. test sensitivity), contacts receive testing, contacts receive a true positive test result. Quality of contact tracing aggregates the probability of: tracers contact a positive index case, a positive index case names contacts, and tracers contact the contacts of the index case. This shows that if testing and contact tracing are done perfectly, we can expect to identify all contacts of infected individuals (illustrated through the colors of the heat map). The blue circles correspond to our simulations using RAT while the white circles correspond to PCR testing use. The two circles in the lower left hand corner correspond to our United States estimates while the two in the upper half correspond to our simulations using estimates from South Korea and Taiwan, where there was stricter contact tracing.

**Figure 3.**
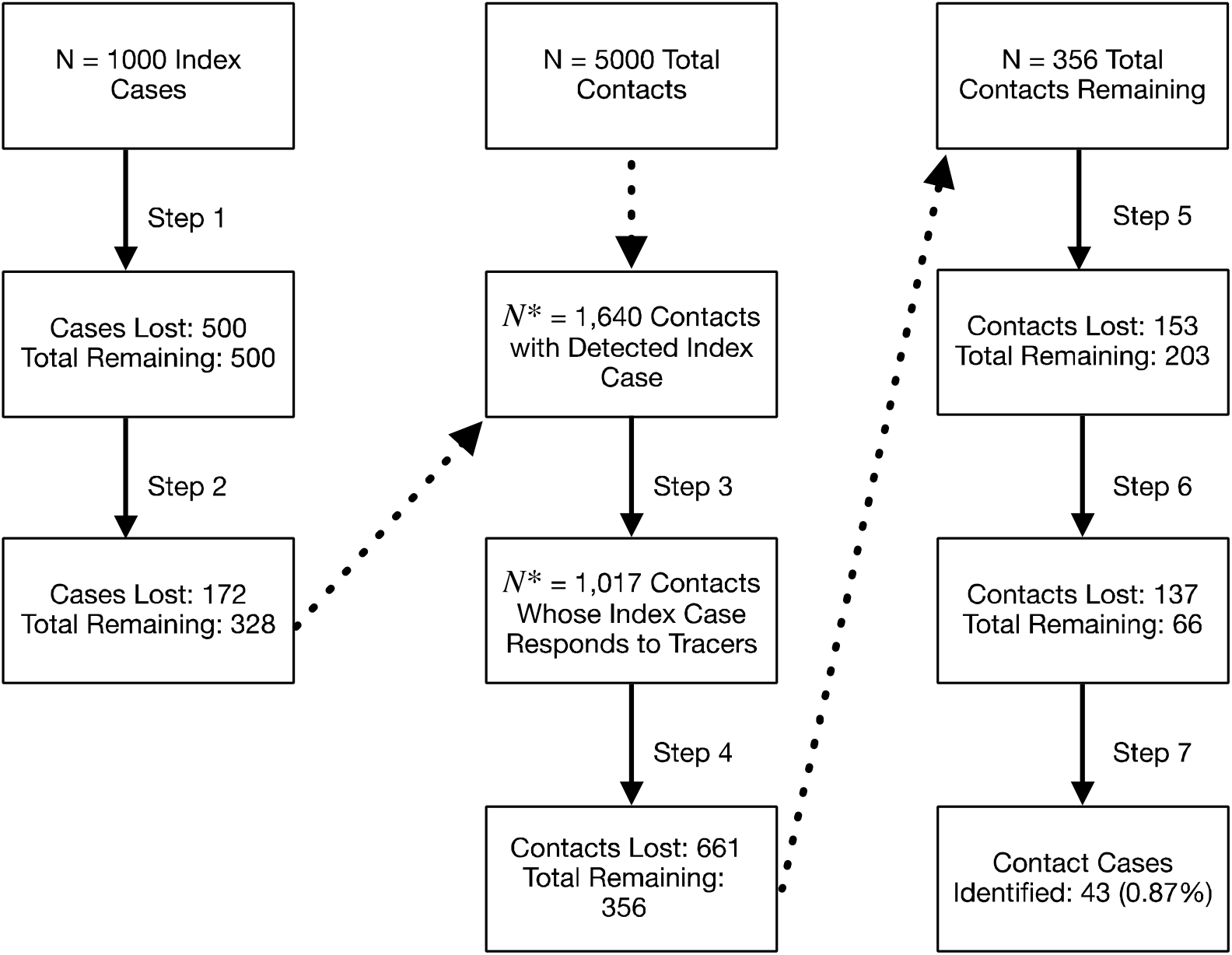
Illustration of Model results. We assume a hypothetical population of 1,000 infected people in a community with 5 unique infected contacts (no shared contacts). The figure depicts the results of our model, tracking the number of losses at each step. We split the tracking of ‘cases’ by first identifying how many index cases were identified. Then, with that information, we moved to correctly identifying positive contacts of those index cases. This example shows that if we had 1,000 people who infected 5 people each (5000 total infected contacts) and assuming the use of RAT, we would expect to correctly identify about 43 of the 5000 secondary cases.

When an index case has been identified, we remove the associated model steps. In our U.S. based example, we estimate a 2.62% (95% uncertainty interval: 2.59%-2.65%) and 3.62% (95% uncertainty interval: 3.58%-3.66%) chance of identifying a positive contact of a known index case using RAT and PCR testing, respectively. In our idealized tracing setting, these increase to 53.3% (95% uncertainty interval: 53.2%-53.4%) for RAT and 73.3% (95% uncertainty interval: 73.2%-73.4%) for PCR testing.

## Discussion

Since its emergence in humans more than three years ago, SARS-CoV-2 has overwhelmed public health institutions globally. The virus still exerts an enormous mortality and morbidity burden worldwide, with nearly 7 million reported deaths thus far [51]. Underlying this is the systematic failure of contact tracing, which was abandoned by most states in the United States by early 2022 [19] and contraindicated in CDC guidance for communities with “sustained ongoing transmission” of COVID-19 [21]. The Lancet Commission report on COVID-19 [52] and several reviews [20,53,54] have highlighted this shortcoming in pandemic response.

Our work takes this a step further by quantifying the extent to which contact tracing failed to identify transmission events in the U.S. and in an idealized setting base on East Asian data. Our demonstrate that the voluntary steps in contact tracing, i.e. seeking testing and interacting with contact tracers, reduced the efficiency dramatically, with fewer than 2% of transmission events identified compared to 62.7% in a setting where testing and compliance with tracing was higher.

Contact tracing formed the basis of modern epidemiological practice, dating back to the investigation of the 1854 Broad Street Cholera outbreak in Britain [55] that led to both a mechanistic understanding of cholera transmission [56] and successful control of the outbreak. A more recent example of highly successful and proactive contact tracing by public health authorities was the effective suppression of monkeypox in the Western US in 2003 [57]. Notably, during the current pandemic, many other countries (such as China, Japan, South Korea, Taiwan, Vietnam and Singapore) were successful at implementing contact tracing in the first two years of the pandemic [14–18].

The impact of poor contact tracing in the U.S. has undermined our understanding of the transmission potential of SARS-CoV-2. For example, the argument that schools do not contribute to SARS-CoV-2 transmission was based in part on the lack of detection of transmission chains in a school setting. Numerous publications showed a lack of contact-traced chains of transmission in a school setting [58–62], while in effect lacking a positive control for the ability to identify onward chains of transmission [63]. It is now clear that SARS-CoV-2 is readily transmitted in schools [64–72], particularly when robust mitigation measures are not in place [73,74]. Indeed, dramatic increases in case detection rates have been observed in studies that relied on surveillance testing, rather than contact tracing [70]. Additionally, this led to the conclusion that the most common source of transmission was gatherings in the home, but it is unclear if this is a consequence of household contacts being easiest to identify or a result of many transmission studies being conducted in settings with strict shut downs, where households were one of the few places where transmission could occur [75]. Also, reports from the West have pointed to a lack of detected transmission chains in air travel [76–78]. These reports are contradicted by careful contact tracing studies from other countries, which have clearly demonstrated person-to-person transmission in flight [79–81], even when robust mitigation measures were in place [82].

Our work has several limitations. We have assumed instantaneous contact tracing, ignoring the impact of tracing delays on infection control which has a significant impact on contact tracing effectiveness against transmission [83,84] due to short incubation period of SARS-CoV-2 [85]. Instead, our estimates for contact tracing effectiveness apply to the informativeness of contact tracing studies, and they form an upper bound for the effectiveness of contact tracing as a transmission prevention measure. We do not account for asymptomatic transmission, again making our estimates an upper bound. The percent of asymptomatic COVID-19 cases is estimated to be anywhere between 1.6% and 56.5% [90–96], with a these cases having a relative reduced infectiousness of 0 to 62% [90–97]. This would mean our estimated 1.65% of transmission pairs identified with PCR testing could be as low as 0.9%, assuming no asymptomatic index cases are identified. We also have not accounted for superspreading, which has been estimated to be a significant feature in COVID-19 transmission [94–96]. This implies that missing a superspreading index case would have tremendous impact on downstream contact tracing efforts and that there is significant stochasticity [97]. We only consider the probability of identifying infected contacts, but it is ideal to also identify uninfected contacts accurately. Finally, we estimate the probability of naming an infected contact using data describing the probability of naming any contacts at all (rather than the probability of naming any given contact). This means that our final estimated probabilities are upper bounds of the true values. Despite representing an upper bound, our contact tracing estimates suggest that U.S. contact tracing studies fail to identify the vast majority of transmission pairs and onward transmission events. This severely limits the inferences that can be drawn from such studies.

Our work points to several key lessons for future public health efforts. First, compliance is a key driver of contact tracing effectiveness. Methods to improve compliance will be crucial for future contact tracing efforts-whether using technological approaches (such as mobile phone or surveillance-camera based tracing) or by making changes to the legal framework around public health efforts (see Supplementary Information S2 for a more on this topic).

Second, there is a pressing need for innovation, to develop contact tracing methodologies that are more resistant to noncompliance. One such approach may be backward contact tracing, which seeks to identify who infected the detected case. Here when contact tracing is executed backward to identify the source of infection (parent), the more offspring (infections) a parent has produced, the more frequently the parent shows up as a contact. Model-based analysis suggests that a backwards contact tracing approach does not require sampling a network at such a large scale as forward tracing [98,99] to understand transmission dynamics, and addresses the problem of low compliance. This approach has been proposed by others for COVID-19 [100–102], and has been empirically shown to be effective, particularly in identifying superspreading events [103], however this is unlikely to be as helpful in reducing transmission.

Third, public health responses to future outbreaks must include educate the public about behaviors with health outcomes, including creating a normative framework around contact tracing compliance. Consistent messaging about limiting transmission and contact tracing are key as has been noted by the Lancet Commission [52], among others [104,105]. This could include reframing messaging to reduce stigma that has often been associated with contact tracing [106] and which undermines contact tracing efficacy [107]. During the HIV epidemic, contact tracers emphasized the index case’s personal responsibility towards the health of their sexual partners [1]. It also includes addressing misinformation, which led many to believe that COVID-19 was a “hoax” [108] and public health measures were overreactions [109].

Finally, we have shown that testing availability and accuracy create a critical gap in contact tracing efforts. Considering only the steps for testing accuracy and cases/contacts receiving testing in our model we find that only 12.4% of possible cases could be identified with RAT, the most available testing modality. To effectively manage future outbreaks, tests need to be sensitive, provide rapid results, and be readily available.

The work presented here adds to the growing body of literature [26,110,111] highlighting the poor performance of contact tracing in the West during the ongoing pandemic and suggests practical fixes for this problem, as we have described. In its absence, public health is forced to rely on population-wide measures for disease spread and will not be able to fine-tune its responses to match the situation. If we are to improve our response to the current crisis, or to others in the future, we must improve our ability to deliver this key function.

## Data Availability

All data produced in the present work are contained in the manuscript

https://github.com/Henry-Bayly/ContactTracingMarkovModel

