## Supplement for "Looking under the lamp-post: quantifying the performance of contact tracing in the United States during the SARS-CoV-2 pandemic"

**Supplement S1. Sources used for each parameter in the model.**

| Source | Value |
| --- | --- |
| Jegerlehner et al. [41] | [0.57, 0.73] |

**Table S1**. Rapid Antigen Testing Sensitivity

| Source | Value |
| --- | --- |
| Kortela et al. [40] | [0.88, 0.92] |

**Table S2**. PCR Testing Sensitivity

| Source | Value |
| --- | --- |
| Stargel et al. [99] | 41% |
| Kalyanaraman et al. [100] | 80% |
| Spencer et al. [101] | 57% |
| Lash et al. 1 [24] | 59% |
| Lash et al. 2 [20] | 77% |
| Bonacci et al. [38] | 68.5% |
| Kanu et al. [102] | 67% |
| Shelby et al. [39] | 48% |
| Hood et al. [43] | 82% |
| Miller et al. [103] | 71% |

**Table S3**. Tracers make contact with a positive case

| Source | Value |
| --- | --- |
| Stargel et al. [99] | 45% |
| Sachdev et al. [104] | 51.1% |
| Lash et al. 1 [24] | 33% |
| Lash et al. 2 [20] | 52% |
| Bonacci et al. [38] | 37% |
| Kanu et al. [102] | 17% |
| Miller et al. [103] | 27% |

**Table S4**. Positive case names contacts

| Source | Value |
| --- | --- |
| Stargel et al. [99] | 58% |
| Sachdev et al. [104] | 45% |
| Lash et al. 1 [24] | 71% |
| Lash et al. 2 [20] | 73% |
| Kanu et al. [102] | 31% |
| Spencer et al. [101] | 55% |
| Miller et al. [103] | 85% |
| Shelby et al. [39] | 28% |

**Table S5**. Tracers make contact with contacts of positive case

| Source | Value |
| --- | --- |
| Stargel et al. [99] | 31% |
| Sachdev et al. [104] | 26% |
| Jones et al. [105] | 45% |
| Matthias et al. [106] | 19.3% |
| Atherstone et al. [107] | 21.3% |
| Sachdev et al. 2 [108] | 40% |

**Table S6**. Contacts of positive case get tested

**Supplement S2. Improving Compliance with Contact Tracing: Approaches And Challenges**

Improved compliance with contact tracing efforts is vital for the effective management of future outbreaks and pandemics. The failed public health response for COVID-19 has normalized a set of behaviors that will serve as obstacles for future public health efforts for disease control. In this section, we discuss ways in which compliance with contact tracing can be improved and challenges that may be faced.

At its simplest, contact tracing efforts can be made mandatory using a legal framework. This can be accomplished by enacting new regulations- for example, by making contact tracing interviews compulsory and subject to perjury penalties [1]. This approach is not likely to be scalable or timely, however. A reliance on large, field-based epidemiological investigations is also unrealistic, given that it would require hundreds of thousands of case workers [2].

An alternative approach is to support contact tracing with software-based approaches, as has been done in countries in the Asia/Pacific region. A number of Asian countries were highly successful at mitigating disease spread in the early part of the pandemic, based on electronic methods that were efficient at identifying contacts using global positioning data from mobile phones, close-circuit television data and credit card transactions [3–5]. For example, Singapore had app-based contact tracing that was optimized for rapid responsiveness while still maintaining a high degree of anonymity [5]. Mandatory electronic methods for contact tracing can streamline the process, with the added benefit of rapidly alerting people when they are exposed to SARS-CoV-2, however these are most effective when implemented in conjunction with traditional public health efforts [6,7]. In the United States, Apple and Google jointly developed contact tracing functionality on their smartphones in the spring of 2020 [8]; however, this app was opt-in and had poor functionality [9] and low adoption rates [10], effectively negating its utility as a public health tool. (A similar lack of effectiveness for voluntary contact-tracing apps has been noted in other countries, such as Iceland [11]).

Mandatory digital approaches are likely to face cultural resistance in the United States, in large part due to several misconceptions in the public domain. There is a perception that the use of contact tracing apps will negatively impact privacy [1,12]. The experience in other countries has shown that this does not have to be the case- properly designed apps can maintain anonymity while still providing the necessary contact-tracing data [13], and legal safeguards (sunsets around data collection, deletion deadlines, access limits and audits [1]) can be put in place. While hacking and misuse of data remains a concern, even in the absence of contact tracing protocols, there is limited electronic privacy in the United States; owning smartphones or Internet of Things (IoT) devices renders the average citizen vulnerable to massive intrusions of privacy from state or non-state actors [14–17]. The United States constitution does not enshrine privacy as a fundamental right, and the question of whether it implicitly protects privacy is controversial [18,19]. Even constitutionally guaranteed fundamental rights are not absolute and are frequently impinged upon by state and federal governments in the context of a compelling state interest, such as protection of citizens’ health [18,20]

Further confusion is caused by the conflation of medical privacy laws with the individual “right to infect others”. Criminal and civil liabilities (up to misdemeanor charges in some jurisdictions) can result from knowingly infecting others with diseases (such as HIV or other STDs [21,22].) Federal legal code also empowers the Surgeon General to make and enforce regulations specifically to prevent the spread of disease [23]. It thus stands to reason that a right to infect others does not exist. An argument has also been made that the use of automated contact tracing is incompatible with personal liberty, from a libertarian perspective [24]. This argument contradicts the guiding principles of libertarian theory, as laid out by the political philosopher John Stuart Mill in his work *On Liberty*: “(T)he only purpose for which power can be rightfully exercised over any member of a civilized community, against his will, is to prevent harm to others” [25].

Any effort to improve compliance for contact tracing must be supported by education. A salient feature of the failed public health response for COVID-19 was the inability of public health bodies to build a shared understanding of the need for public health responses in the first place. In the context of COVID-19, misinformation about COVID-19 led to a range of reactive behaviors around noncompliance (such as a lack of compliance with masking and quarantine requirements). Such behaviors, while individually rational, effectively undermined the ability to limit COVID-19 spread (a phenomenon that was predicted by us very early in the pandemic [26].) In response to noncompliance, public health bodies sought to “meet the public where they are” by changing guidelines to normalize dysfunctional behaviors (for example, relaxing requirements for masking and quarantine [27–30].) In doing so, the public health response shifted the responsibility for disease control on to the individual. Individual public health measures are both oxymoronic and infeasible during a pandemic.

To prevent such failures from occurring again in the future, public health efforts should seek to create a normative framework around the right behaviors, starting from accepting the fact that unhealthy behaviors exist and carefully exhibiting why they are harmful. Thus, any future public health response to outbreaks or pandemics must include a component of educating the public about those behaviors that will lead to better outcomes for the public itself. The WHO, in its Shanghai Declaration [31] on promoting health in the 2030 Agenda for Sustainable Development [32] has drawn attention to the importance of health education methods and strategies for Low and Middle Income Countries. Because the burden of infectious disease falls disproportionately on lower-income communities, better communication around public health outcomes addresses issues of inequity within the United States as well. Consistent messaging around the need for limiting transmission and the need for contact tracing are key in building -or rebuilding- trust.

Restoring contact tracing functionality is a crucial first step in rebuilding the capability of public health organisations to respond effectively to future outbreaks. Central to that effort will be addressing the issue of compliance with contact tracing efforts- by no means an easy challenge.
